# The Application of Trimetazidine in Healthy Individuals: A Systematic Review

**DOI:** 10.1101/2022.05.09.22274810

**Authors:** Eduard Bezuglov, Maria Shoshorina, Andrey Zholinsky, Zbigniew Waśkiewicz, Anton Emanov, Ryland Morgans, Vladislav Bertholz, Vasilisa Zelenskaya, Artemii Lazarev, Maria Sokolskaya, Oleg Talibov

**Author notes:** **Corresponding author:** Maria Shoshorina.

## Abstract

**Background:** Currently, there is no evidence to support TMZ administration aiming to enhance any parameter of physical performance or post-exercise recovery in healthy individuals of the general population and athletes.

**Objective:** Considering the lack of empirical data, from a scientific and practical perspective it would be interesting to identify research with high methodological quality that examines the effects of trimetazidine (TMZ) on healthy individuals of the general population and athletes of varying age.

**Methods:** Data sources included English articles that were searched by keywords in the PubMed and Scopus databases and the Cochrane Library and published prior to November 2022. Thus, a systematic review of the scientific literature was performed utilizing databases with a traditional PRISMA methodology. An initial search by keyword found 2673 publications and the screening process selected 66 articles, of which two articles met the inclusion criteria.

**Results:** Two trials examining the effect of TMZ on healthy members of the general population that were published in 2017 and 2019 were analyzed. Athletes were not recruited for this type of research.

**Conclusions:** Currently there are no data reporting a positive effect of TMZ on physical performance, post-exercise recovery, or other health parameters in members of the general population, while its administration is associated with the development of relatively common adverse effects.

## Introduction

Modern elite sport places high demands on the human body, promoting the regular introduction of new methods and substances aiming to improve physical performance and post-exercise recovery. One of the most commonly employed methods to improve athletic performance is the administration of various pharmacological substances, biologically active supplements, and nutraceuticals. Various data suggest that the majority (40-100%) of elite athletes use or have used biologically active food supplements [1, 32, 11]. Moreover, elite athletes use food supplements more often than their non-elite colleagues [17]. The average number of used food supplements is 1.7 to 3.4 per single athlete throughout the competition season [1, 29]. Currently there is a number of proven substances that positively effect various aspects of physical performance, however, there is an association with health risks for users.

This primarily refers to methods and substances included in the World Anti-Doping Agency (WADA) Prohibited List, the organization established in 1999 and aimed at providing doping-free participation in competitions. To be included on the prohibited list, a method or substance must satisfy a minimum of two from three criteria:

- potential enhancement of sport performance;
- potential health risk;
- violation of sporting spirit.

At present WADA Prohibited List contains over 20 classes of prohibited methods and substances, while only six are proven to positively affect physical performance. These are anabolic agents, growth hormones, stimulators, beta-agonists, androgen receptor modulators, and some metabolic drugs [33]. According to Heuberger et al., TMZ is one of the substances that has no proven efficiency in healthy individuals although is included in Hormones and Metabolic Modulators class S4, which is mainly prescribed for managing angina [14]. Trimetazidine was originally developed in 1963 by Servier Company, although nowadays there are more than 30 commercial names for TMZ drugs. The TMZ drug is produced in capsules or tablets with an instant or extended release. The quantity of TMZ per one drug unit varies from 15 to 80 mg.

Trimetazidine action implies partial inhibition of beta-oxidation of fatty acids by selective inhibition of 3-ketoacyl-CoA thiolase enzyme in cardiomyocytic mitochondria, resulting in the intensification of glucose oxidation and acceleration of glycolysis with glucose oxidation. This mechanism is thought to protect myocardium from ischemia and cell damage by the products of lipid peroxidation [16, 9]. In the majority of countries TMZ is used to stabilize angina as a combination therapy and secondary treatment [5]. In other countries TMZ is also registered to treat chorioretinal vascular disorders and tinnitus [24]. It should be noted that these indications were withdrawn by the Ministry of Health for France in 2011 and by the European Medicines Agency in 2012 due to a poor tolerability profile [23].

Despite TMZ being known for decades, it has only been named as a prohibited competition substance (Stimulators class S6) since 1^st^ January 2014 and banned since 1^st^ January 2015 (Hormones and Metabolic Modulators class S4). The first athletes suspended for TMZ usage were Ukrainian skier Marina Lisogor and Chinese swimmer Sun Yang, i.e. cyclic sport athletes. Additionally, in recently published scientific literature (prior to 2020), no study demonstrated the efficiency of TMZ on any aspect of physical performance or post-exercise recovery in healthy individuals of the general population [14, 3].

Considering the lack of empirical data, from a scientific and practical perspective it would be interesting to examine research with high methodological quality that assesses the influence of TMZ in healthy members of the general population and athletes of different levels. A further aim of the study is to determine the range and rate of adverse effects after TMZ administration.

## Materials and methods

The scientific literature search was conducted utilizing the Pubmed and Scopus databases and the Cochrane Library to identify trials addressing the effects of TMZ on healthy subjects. The search was conducted in accordance with PRISMA guidelines.

Literature screening was performed in November 2022 using the following search request: (“Trimetazidine Dichlorhydrate” OR “Trimetazidine Dihydrochloride” OR “Trimetazidine Hydrochloride” OR “Trimetazidine”) AND (athletics OR sports OR human OR animal OR endurance OR strength OR speed OR coordination OR “cognitive functions” OR recovery OR hypoxia OR contamination OR wada OR doping OR training OR competitions OR performance OR “adverse event” OR “side effect” OR efficacy OR effectiveness OR adolescents OR children OR “extreme ambient conditions” OR “military medicine” OR “military personnel” OR “Mixed martial art”) AND NOT (surgery OR angina OR infarction OR tinnitus OR “heart failure” OR combination). All discovered and eligible articles were examined. To formulate eligibility criteria PICOS was used. The inclusion criteria were:

1. The article is a clinical study;
2. The subjects of the study were healthy individuals;
3. The research focused on the effects of TMZ of trimetazidine on the body;
4. The intervention was the use of trimetazidine versus placebo.

In addition, the selection of articles was based on a detailed review of studies in which:

1. The intervention was carried out on animals;
2. The determination of trimetazidine in the blood and urine of healthy volunteers was studied;
3. The side effects of trimetazidine were described.

All identified studies have been assessed for risk of bias using the Revised Cochrane risk-of-bias tool for randomized trials (RoB 2) [27]. Cases of disagreement in assessments of the risk of bias between the reviewers were resolved by discussion or with consultation with a third reviewer if needed.

## Results

Keyword searches identified 2673 publications. Twelve articles were excluded as the literature was not in English language. Title and abstract screening selected 66 articles that were thoroughly examined for eligibility using the following criteria (Figure 1.)

**Figure 1.**
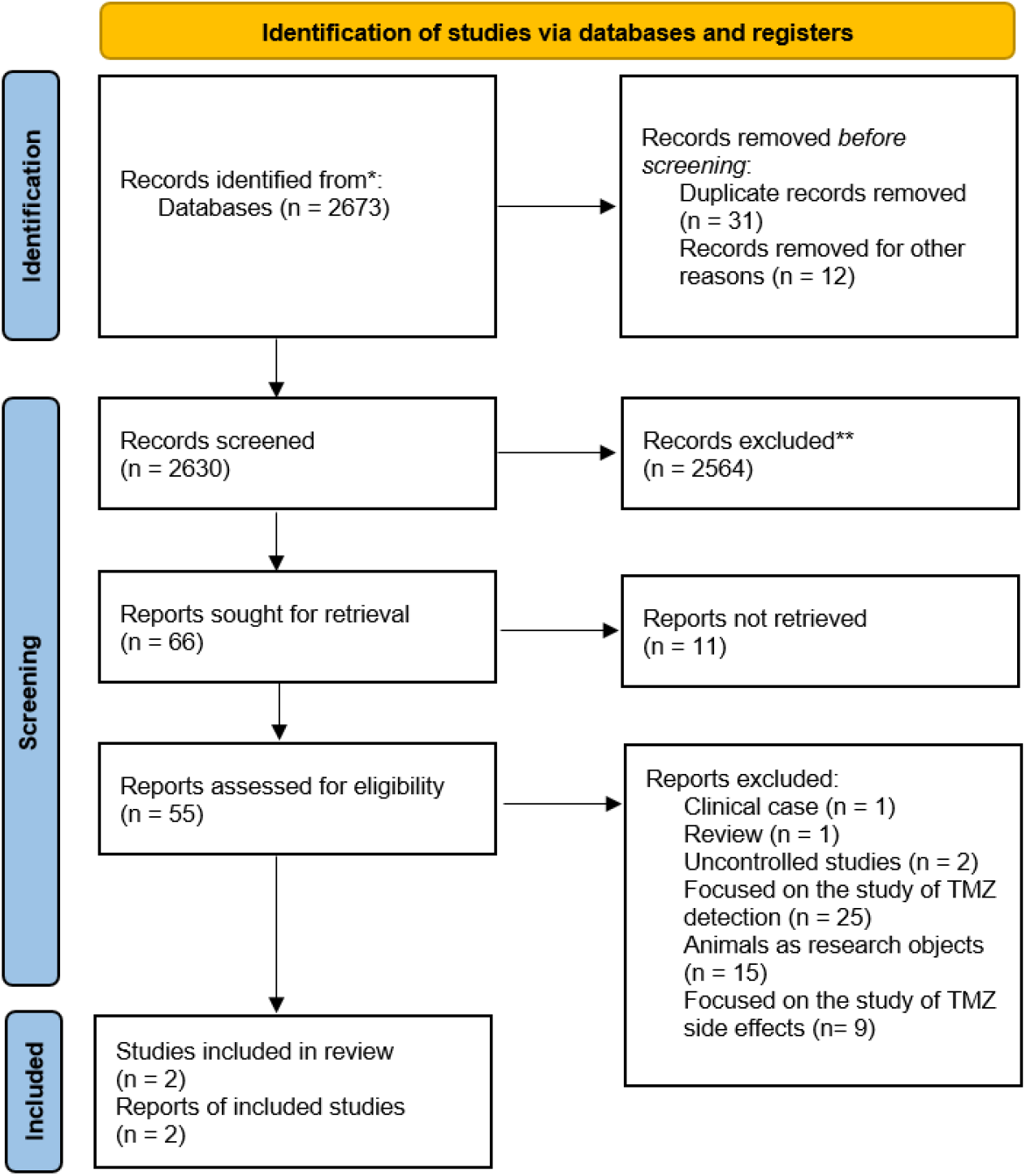
Selection of studies.

**Figure 2.**
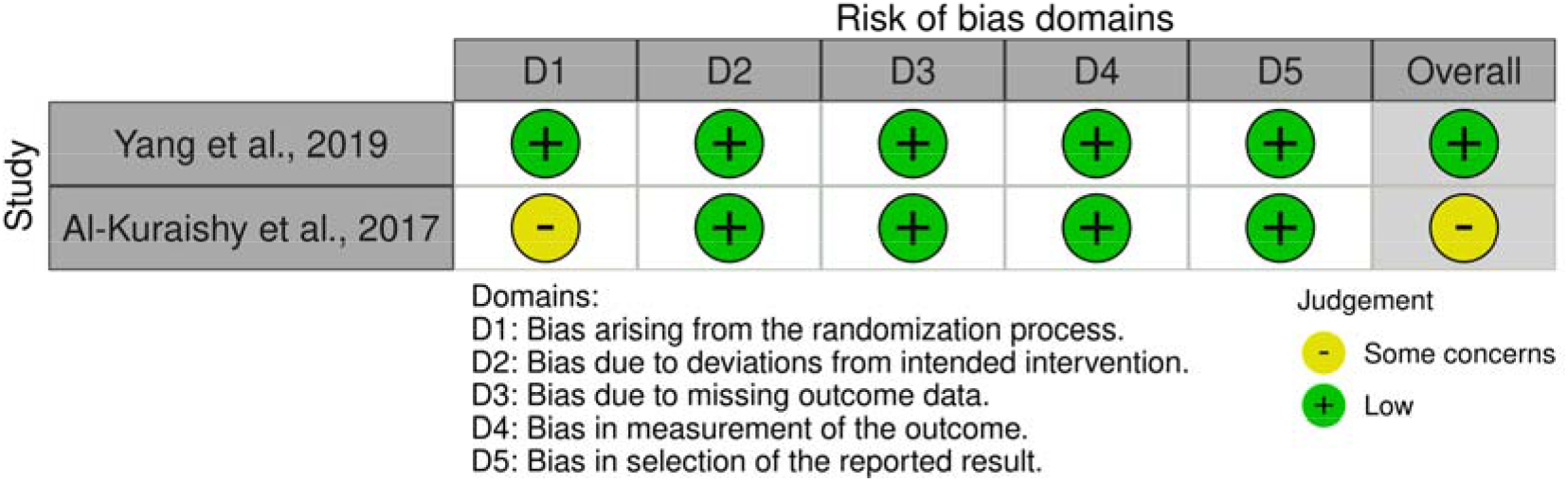
Risk of bias in the studies included.

The excluded articles are listed above one clinical case, one literature review and two uncontrolled studies, 25 articles dedicated to the methods of TMZ detection rather than its effects, 15 articles used animals as subjects and 9 articles assessed TMZ safety and tolerability.

In summary, only two randomized controlled trials (RCTs) examined the effect of TMZ on healthy volunteers who were not elite athletes, published in 2017 and 2019 [2, 34], i.e., studies of high methodological quality (Table 1).

**Table 1.**
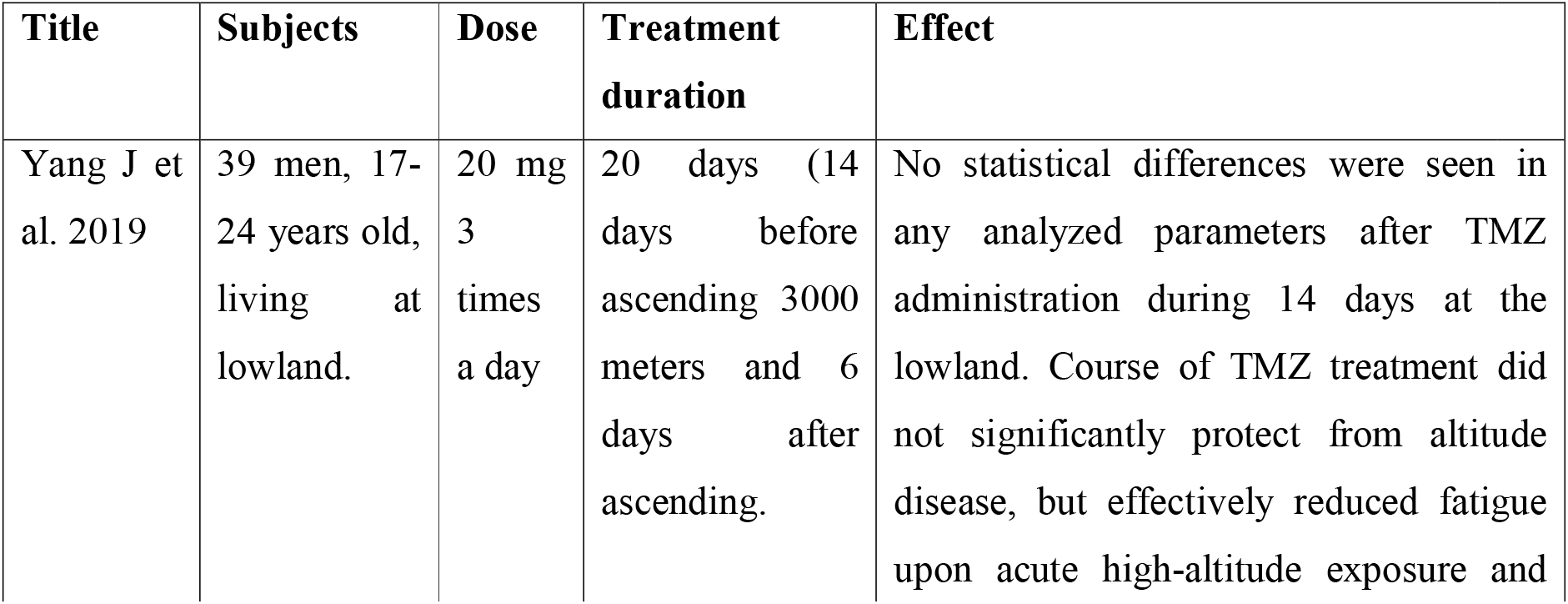
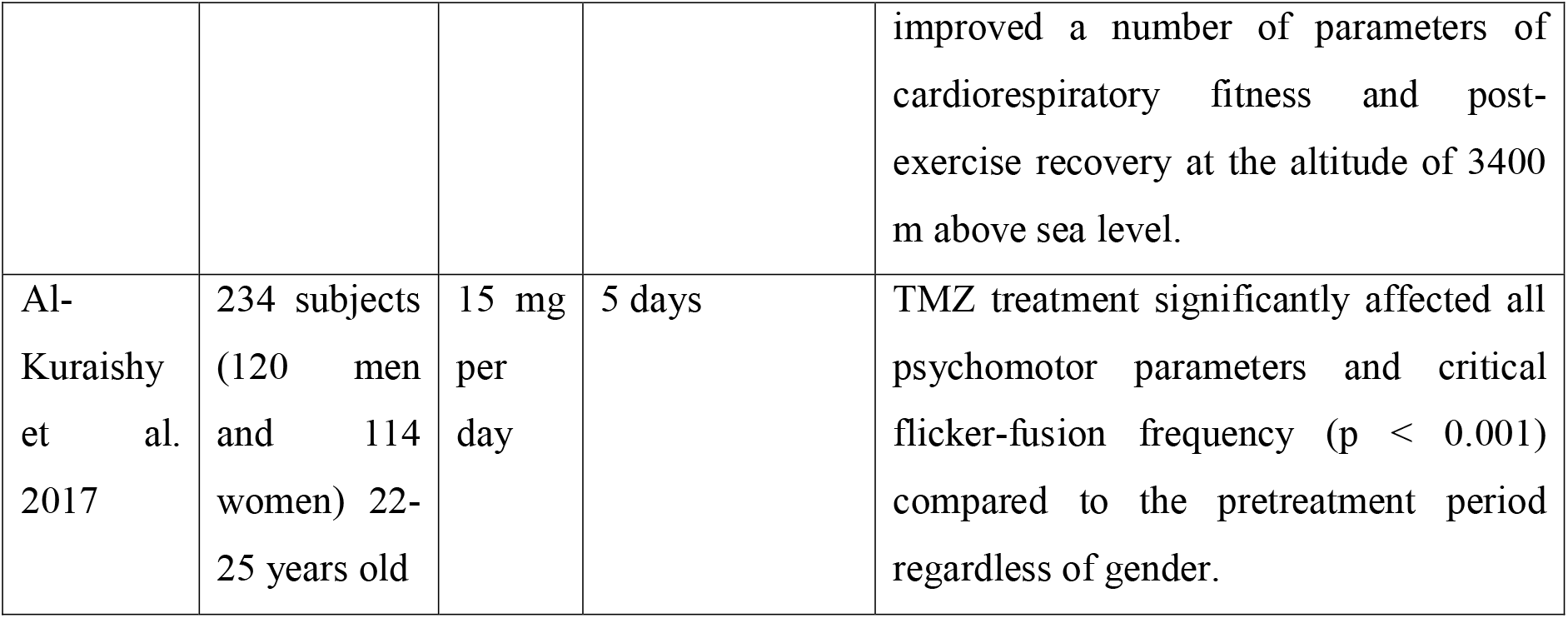
Studies investigating the effect of trimetazidine use in healthy volunteers.

| Title | Subjects | Dose | Treatment duration | Effect |
| --- | --- | --- | --- | --- |
| Yang J et al. 2019 | 39 men, 17-24 years old, living at lowland. | 20 mg 3 times a day | 20 days (14 days before ascending 3000 meters and 6 days after ascending. | No statistical differences were seen in any analyzed parameters after TMZ administration during 14 days at the lowland. Course of TMZ treatment did not significantly protect from altitude disease, but effectively reduced fatigue upon acute high-altitude exposure and improved a number of parameters of cardiorespiratory fitness and post-exercise recovery at the altitude of 3400 m above sea level. |
| Al-Kuraishy et al. 2017 | 234 subjects (120 men and 114 women) 22-25 years old | 15 mg per day | 5 days | TMZ treatment significantly affected all psychomotor parameters and critical flicker-fusion frequency ($p < 0.001$) compared to the pretreatment period regardless of gender. |

From the analysis performed, the risk of bias in the study by Yang J. et al. was low and the study by Al-Kuraishy et al. had some concerns, because of poorly described background information about the participants, which makes it difficult to assess the quality of the randomisation.

## Discussion

Our results identified only two studies of TMZ in healthy individuals representative of the general population [2, 34]. In this studies the time period for TMZ usage was short (5 and 20 days). No studies investigated individuals in specific conditions (elite athletes, military personnel, ballet dancers, etc.), suggesting the lack of TMZ support in such environments.

Yang et al. examined 39 randomized volunteers administering 20 mg of TMZ or a placebo three times a day (20 and 19 subjects, respectively) for two weeks post-treatment and examined a number of cardiorespiratory parameters. Following the acute shift in altitude (400 m above sea level to 3400 m above sea level without acclimatization) no inter-group differences were found, although the cardiorespiratory parameters in the TMZ group significantly passed the parameters of the placebo group. Despite these clear differences, these data poorly apply to athletic activities with the possible exception of mountain sports. This may be partly explained and associated with the climatic condition of altitude (3400 m in the Yang et al., study), as no training or competitions are organized in such conditions except for certain extreme sports. Furthermore, if training and highland living at an altitude of 2000-2500 m is included in the preparation programs of athletes with the aim of creating conditions for the pre-conditioning of muscles, cardiovascular system, and blood under relative oxygen deficiency this may prove beneficial. Furthermore, TMZ application reduces the contribution of relative hypoxia and alleviates the altitude effects [34].

The second study performed in healthy individuals was not directly related to the analysis of functional parameters and could not serve as reliable source of information regarding TMZ efficiency. Al-Kuraishy HM et al, 2017, determined the influence of five-days of TMZ administration on psycho-motor reactions. The authors demonstrated that such a dosing regimen may improve psycho-motor performance and vigilance in normal healthy volunteers through improving reaction time and critical flicker-fusion frequency [2].

Two uncontrolled studies published in 1985 and 1993 were also analysed in detail. In the study by Maridonneau-Parini I et al, 1985, seven volunteers consumed TMZ for seven days and showed a decrease in indices characterizing the severity of erythrocyte damage by peroxidation products. However, no functional parameters such as several parameters named here were assessed [18]. Devynck MA et al., 1993, supplemented blood previously taken from healthy volunteers and patients with arterial hypertension with TMZ. *In vitro* TMZ effects were shown in ADP-induced platelet aggregation and adenylyl cyclase activity, which did not represent the functional parameters analyzed [6].

An important issue for administering any pharmacological substance is safety and tolerability. According to FDA guidelines, all side effects can be classified as: very common, common, uncommon, rare, and very rare. The adverse effects rate is between 1 and 10 cases per 100 administrations and this is considered common. Our research only examined trials of high methodological quality reporting the rate of adverse effects in clinical administration of TMZ in patients with cardiovascular diseases. Three of the largest trials assessing the efficiency of TMZ (2026 patients) reported only 5% (101 patients) with unfavorable effects. The most common side effect described included nausea or vomiting (1.73%), dry mouth or hot flushes (0.74%), headache (0.64%), diarrhea or distension (0.35%), sleep disturbances (0.30%), and weakness or fatigue (0.25%) [5]. Further studies reported parkinsons, tremor, and gait impairment during TMZ administration [4, 19, 21].

The safety of any food supplement and medication used by elite athletes against un-intentional doping is crucial. Since TMZ is a prohibited substance and its detection in doping tests is followed by long-term suspension of up to four years, it is a substantial problem. According to available data, contamination of biologically active supplements and nutraceuticals with prohibited substances not listed on the label is common and varies from 12% to 58% [20]. Duiven et al. demonstrated that 38% of food supplements bought online (21 brands in 17 online shops) contained undeclared banned substances, including consistently prohibited items by WADA, such as anabolic steroids and metabolic modulators, which if found in a tested sample is punishable by a two-to-four-year suspension [7]. Contamination of any food product or WADA-accredited medication with prohibited substances is possible. However, unlike the established data on the contamination prevalence of biologically active supplements, information regarding the contamination of food and permitted drugs with prohibited substances resulting in Adverse Analytical Findings (AAFs) is scarce and mostly confined to substances such as anabolic agents (clenbuterol, exogenic anabolic steroids, and SARMs) and diuretics [8, 10, 12, 22, 28, 31]. As for other prohibited substances, only single cases have been described and this quantity is unlikely to grow extensively. For example, cases of food contamination with furosemide and letrozole have been reported previously where athletes escaped suspension or were mitigated by re-assuring the disciplinary board of an un-intentional administration of the prohibited substances [25].

Although, in some cases, official drugs permitted for sportsmen were found to be contaminated with prohibited substances not listed on the label. Helmin et al. described such a case in 2016, where the coating of ibuprofen, a non-steroid anti-inflammatory drug, contained trace amounts of a banned substance, diuretic hydrochlorothiazide, which resulted in disciplinary proceeding. Eventually, the athlete was acquitted [13]. There are currently two documented cases where TMZ contaminated a biologically active supplement and a multi-vitamin complex. In 2016 an American swimmer Madisyn Cox was suspended for two years for doping. The positive TMZ test was taken out-of-competition period. The Hearing Panel of FINA International Swimming Federation upheld the sanction since Cox failed to list a likely source of this substance in her sample. Although, she managed to prove that her vitamin and mineral complex contained TMZ and the Court of Arbitration for Sport reduced her two-year suspension to six months. Furthermore, during the 2018 Olympic Games in Korea, TMZ was discovered in the doping test of Nadezhda Sergeeva, a Russian bobsledder. Her Olympic results were revoke and she was disqualified for eight months. Later she managed to prove that methionine amino acid, an authorized drug she consumed, contained trace amounts of TMZ and thus had unknowingly ingested this prohibited substance. The information concerning the presence of TMZ was not listed in the drug label. Therefore, in October 2018, the Court of Arbitration for Sport confirmed that the athlete received the prohibited substance involuntarily and allowed her to continue competing.

Finally, it should be emphasized that the detection of TMZ in doping tests is currently exclusive and the majority of cases involve athletes from cyclic sports. Thus, according to the 2019 WADA report, the detection of TMZ in doping tests was associated with only five (5 from 362) AAFs. This is only 1% of documented cases referred to the Hormone and Metabolic Modulators class S4, which includes TMZ. Furthermore, such substances of this class such as tamoxifen, meldonium, and clomifene were identified as AAFs in doping tests 80, 79, and 73 times in 2019, respectively [30]. Moreover, it should be mentioned that our search did not detect any trials investigating the influence of TMZ on any health or physical performance parameters in people above 18 years old.

Considering the limited practical application of TMZ even prior to its prohibition (0.1% - 0.23% of total tests analyzed in anti-doping laboratories in Warsaw and Cologne until 2010), it is fair to conclude that TMZ application has always been scant [26, 15]. It is important to note that for athletes who are subjected to regular testing, a course of TMZ treatment is extremely difficult, as athletes have to regularly update the ADAMS system with their location and are always available for testing by the National Anti-Doping Organizations and International Sports Federation. Furthermore, since the TMZ measurement threshold in urine is 0.5 ng/mL in WADA-accredited laboratories and this concentration is detectable for a minimum of five days even after a single use, the intentional TMZ administration by athletes is completely unreasonable and associated with extremely high risk of long-term suspension. Together with the absence of empirical scientific data supporting TMZ efficacy for healthy people as a pharmacological agent to improve exercise tolerance, this allows the conclusion that TMZ administration is inadvisable.

In conclusion, this systematic review provided the following conclusions:

1. currently, there is no evidence to support TMZ administration aiming to enhance any parameter of physical performance or post-exercise recovery in healthy individuals of the general population and athletes;
2. the prevalence of TMZ application, even prior to its prohibition, is scarce and confined to cyclic sport athletes;
3. no published TMZ studies examined children and adolescents under 18 years old;
4. the administration of TMZ may lead to the development of adverse effects (up to 5%) including those associated with impaired co-ordination and loss of movement precision.

## Data Availability

All data produced in the present work are contained in the manuscript

## Acknowledgments

Not applicable.

## Declaration

### Ethics approval and consent to participate

Not applicable.

### Consent for publication

Not applicable.

### Availability of data and materials

The datasets used and/or analysed during the current study are available from the corresponding author on reasonable request.

